# Effect of PCI on Clinical Prognosis of Chronic Coronary Artery Occlusion

**DOI:** 10.1101/2024.06.05.24308526

**Authors:** Lei Jiang, Mei Wang, Pu Liu, Dong Liang, Jiangpeng Wang, Haoyu Meng, Xiangqing Kong, Leilei Chen

## Abstract

**Background:** Coronary chronic total occlusions (CTOs) are considered to increase the risk of adverse clinical outcomes. The purpose of this study was to evaluate whether long-term clinical outcomes could be improved by successful percutaneous coronary intervention (PCI) over optimal medical therapy (OMT) in CTO patients.

**Methods:** 258 consecutive patients with CTO lesions undergoing PCI at the First Affiliated Hospital of Nanjing Medical University from January 2011 to December 2017 were enrolled. After 25 were excluded due to CABG surgery, a total of 233 patients who met the enrollment criteria were divided into successful CTO-PCI group (n=187) and CTO-OMT group (n=46) based on the treatment received. The study primary endpoint was major adverse cardiac cerebrovascular events (MACCE), including cardiac death, recurrent myocardial infarction, unplanned revascularization, and stroke. The secondary endpoint was all-cause death.

**Results:** During a median follow-up of 78 months, PCI treatment significantly improved MACCE incidence survival probability compared with OMT (29.55% vs 21.95% p=0.028). There was no difference between these two groups in secondary endpoint (p=0.93). There was also no significant difference in MACCE between single vessel CTO lesions and single vessel CTO combined with multiple vessel lesions(p=0.54). The cumulative survival of LAD is the highest among different branch lesion groups(p=0.044). Elderly patients (≥65 years) in PCI group showed a significant decrease of MACCE incidence compared with OMT (35.00% vs 21.33% p=0.001).

**Conclusions:** Successful PCI in CTO patients is associated with a significant decrease of MACCE compared with OMT.

## Introduction

Chronic total occlusions (CTOs) of coronary arteries are defined as the complete luminal obstruction of a native coronary artery with an occlusion duration of >3 months[1]. Some studies reported that CTO patients count for incidence of 10% to 30% on diagnostic angiograms[2-5], percutaneous coronary intervention (PCI) of CTO patient is a vital challenge for interventional cardiologists. With the improvement of equipment and technologies, PCI on CTO is growingly pursued, and the success rate is getting higher [6]. However, its efficacy remains controversial[7,8]. Some observational studies comparing CTO-PCI versus CTO-OMT have demonstrated that successful CTO-PCI was associated with better outcomes [9-11], in which the major benefit is the improvement of symptoms. Nevertheless, some randomized controlled trials (RCTs) concluded inconsistently and questioned the benefit of CTO-PCI [7,12-14]. Overall, CTO-PCI remains uncertain on clinical benefits. Therefore, it is necessary for in-depth studies to explore whether successful CTO-PCI revascularization could bring clinical benefits for the CTO patients. The purpose of this retrospective study was to investigate the effect of successful CTO-PCI intervention on the prognosis, and the incidence of major adverse cardiac cerebrovascular events (MACCE), including cardiac death, recurrent myocardial infarction, unplanned revascularization, and stroke.

## 2. Materials and Method

### 2.1 Study Design and Population

This study enrolled 233 patients with at least one coronary CTO lesion at our hospital in between January 2011 and December 2017. Exclusion criteria were: (1) patients with ST-segment elevation myocardial infarction (STEMI); (2) a history of coronary artery bypass grafting (CABG); (3) cardiogenic shock; or (4) malignant tumor. Patients were referred for PCI based on CTO-related symptoms or evidence of viability, or corresponding ischemia in the area of the CTO artery. The enrolled patients were divided into 2 groups, i.e., CTO-PCI group (n=187) and CTO-OMT group (n=46). The strategy to perform PCI on CTO patients was dependent on some factors, including co-morbidity, technical challenges, operators’ preference[15-17]. Detailed data on demographic and clinical characteristics, procedures, complications were documented and reviewed on hospital database. Follow-up data were obtained through hospitalization notes, phone communications or outpatient visits.

### 2.2 Ethics

This was a single-center, retrospective observational study which was approved by the Ethics Committee of the First Affiliated Hospital of Nanjing Medical University. (2023-SR-893)

### 2.3 Study Endpoint and Evaluations

CTO was established as complete coronary occlusion with thrombosis in myocardial infarction (TIMI) grade 0 flow ≥ three months[18]. Major adverse cardiac and cerebrovascular events (MACCE) were defined as the composite of cardiac death, recurrent myocardial infarction, unplanned revascularization, and stroke. The secondary endpoint was all-cause mortality. All-cause death included deaths from any cause. Stoke was defined as a new focal neurological deficit occurred suddenly from cerebrovascular irreversible cause (or resulting in death) within 24 hours and was not led by any other identifiable cause. Technical success referred to CTO revascularization in treated segment with achieving residual stenosis < 30%, and TIMI flow grade 3[19].

### 2.4 Statistical Analysis

Continues variables are presented as mean ± standard deviation (SD), being tested for differences with Student’s T-test, and categorical variables in frequencies and percentages with Pearson’s chi-square test and Fisher’s exact test. The survival curve was estimated and drawn by Kaplan Meier method, and the difference between the two curves was compared by Log-rank method. Cox proportional hazards regression model was used to detect independent risk factors for endpoint events. Hypothesis tests are all bilateral, P<0.05 is statistically significant difference. All data were analyzed by R software (version 4.3.1).

## 3. Result

### 3.1 Baseline clinical characteristics

233 patients were consecutively enrolled in this study and assigned into two groups: 187(80.3%) patients were managed by successful PCI revascularization (CTO-PCI group) and 46 (19.7%) patients who failed in PCI attempt and started medical treatment (CTO-OMT group). The mean age of patients was 62.7±1.9 years old, and male were 84.5%. (1)Baseline patients characteristics of both CTO-PCI group and CTO-OMT group were listed in Table 1. CTO-PCI group had a higher incidence of previous PCI procedures. (2) There was clinical baseline data difference in administration of ACEI/ARB between single vessel CTO lesions and single vessel CTO combined with multiple vessel lesions, which was demonstrated in Table 2. (3) Table 3 demonstrated clinical baseline comparison showing no difference among different vessels. (4) Table 4 showed the clinical baseline comparison between two treatment groups in elderly (≥65 years old), who had prior CTO-PCI treatment.

**Table 1.**
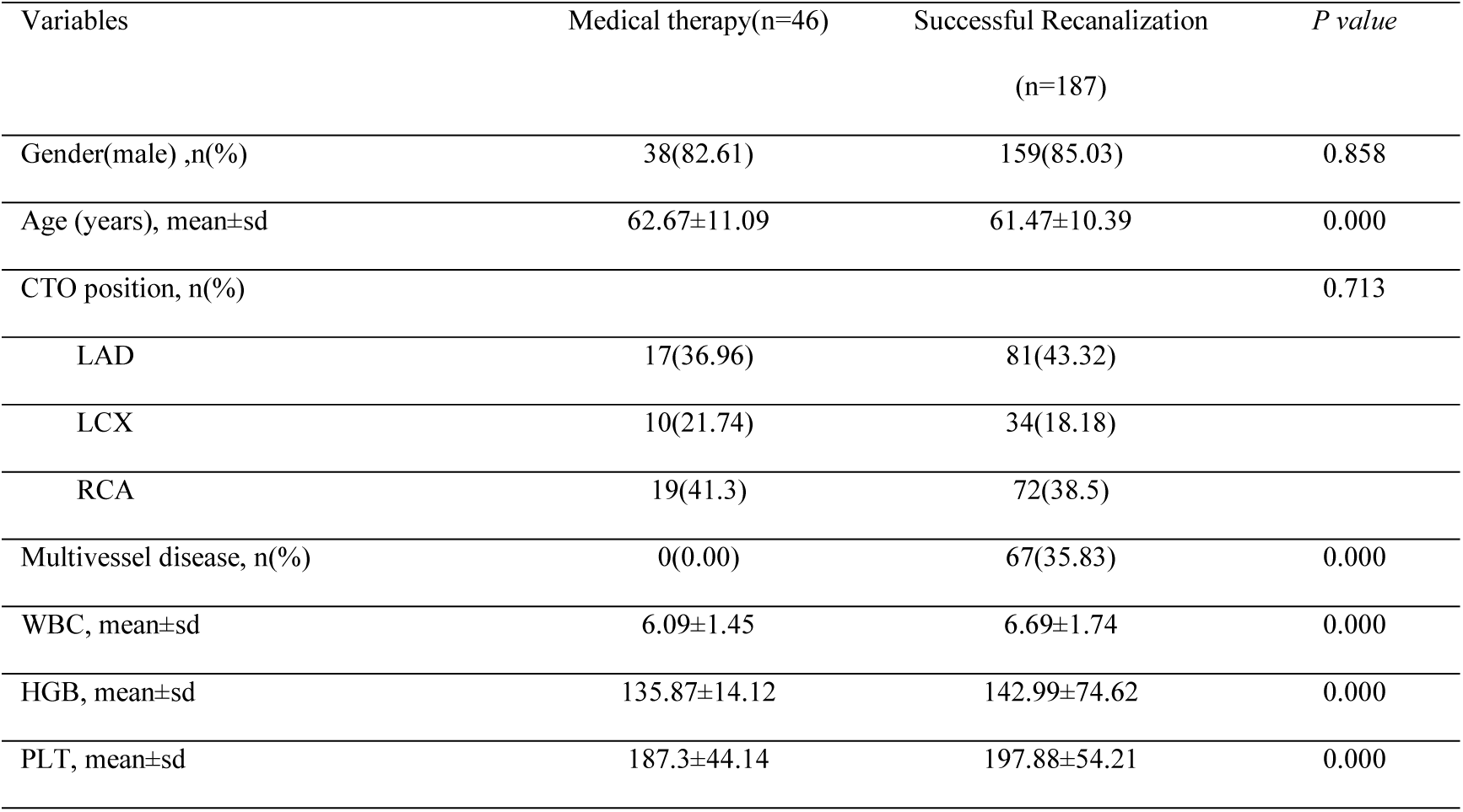
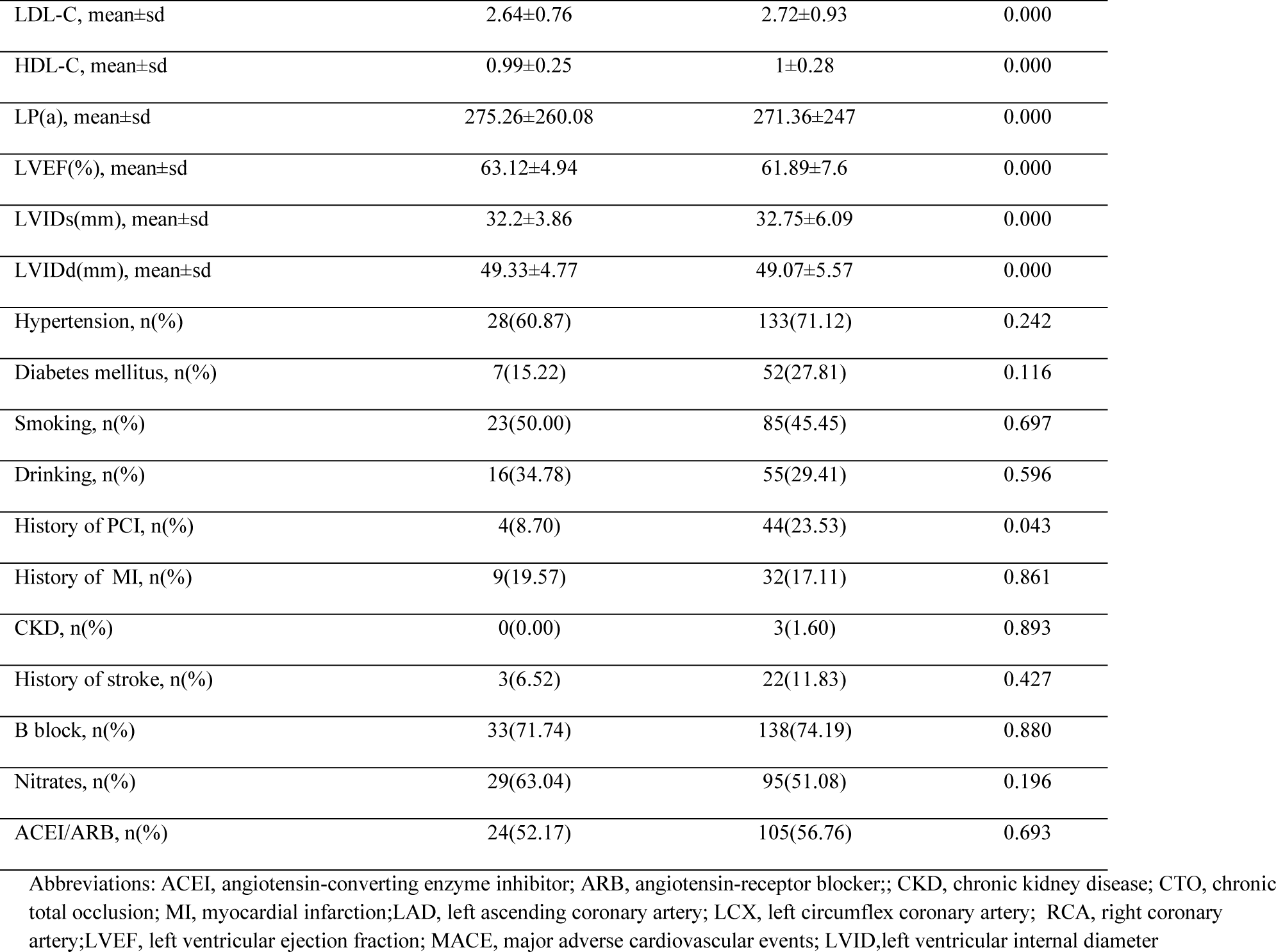
baseline clinical and in-hospital outcome of all patients in the Successful Recanalization and Medical Therapy Groups.

**Table 2.**
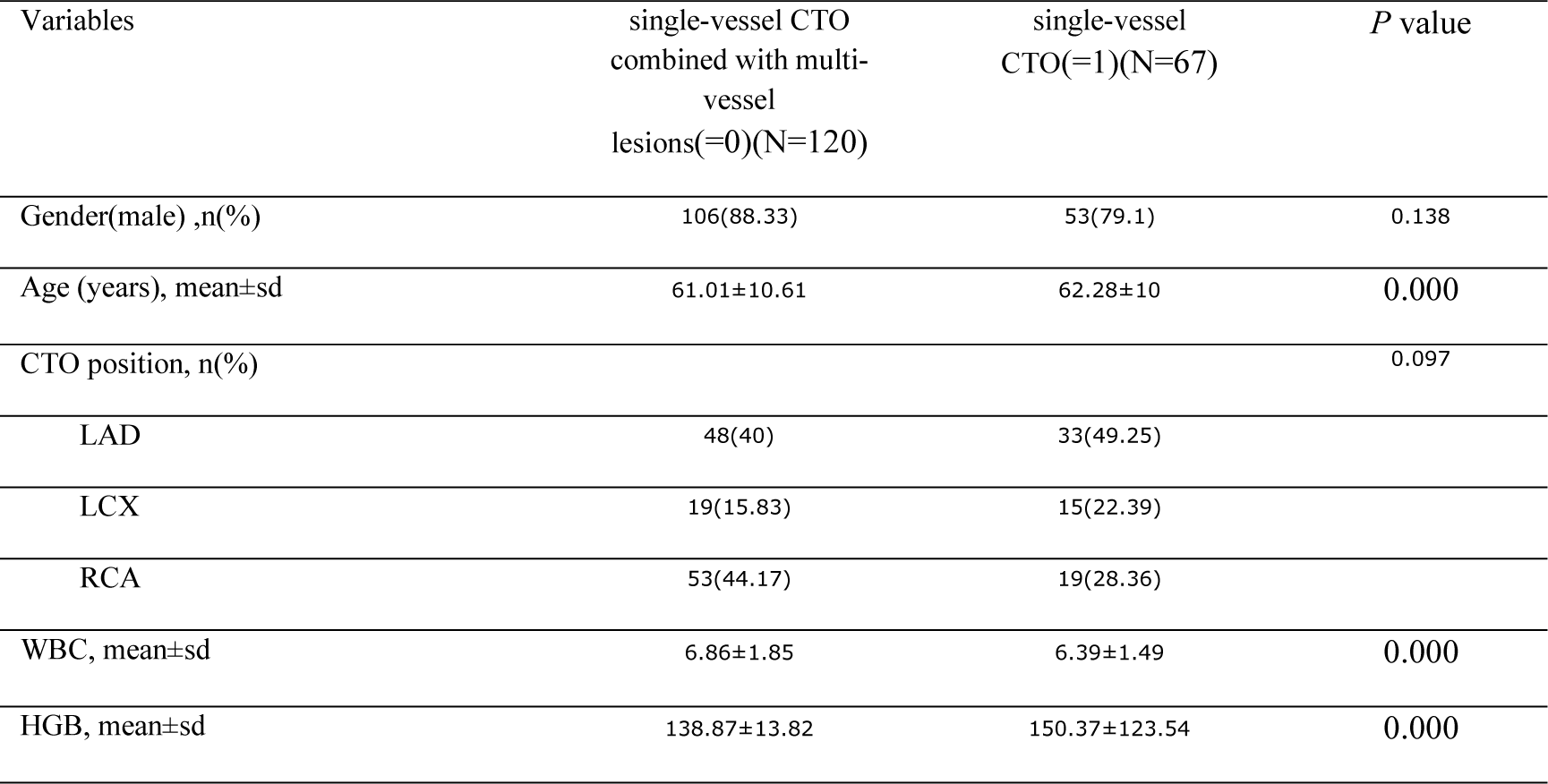
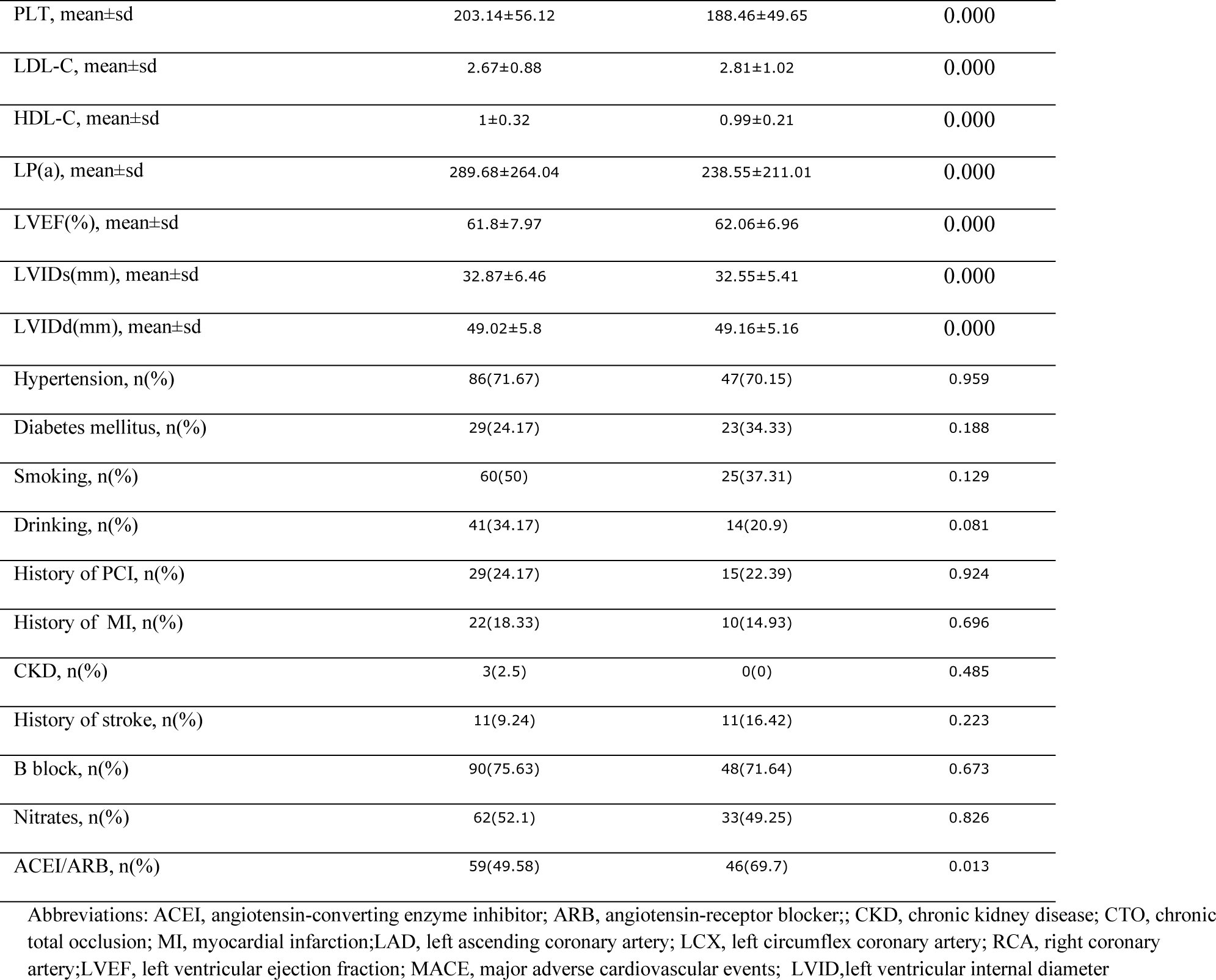
baseline clinical and in-hospital outcome of all patients in the single-vessel CTO and single-vessel CTO combined with multi-vessel lesions.

**Table 3.**
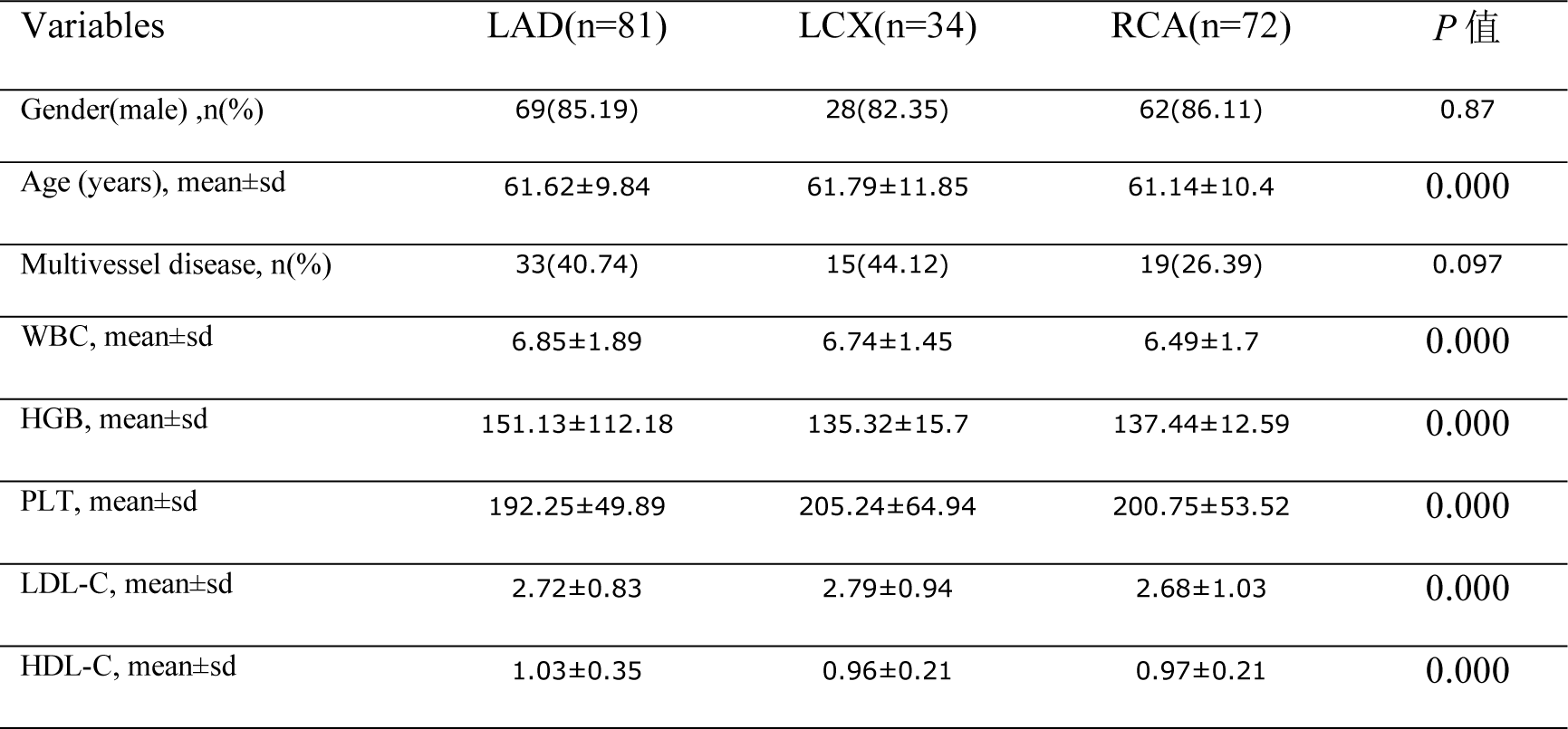
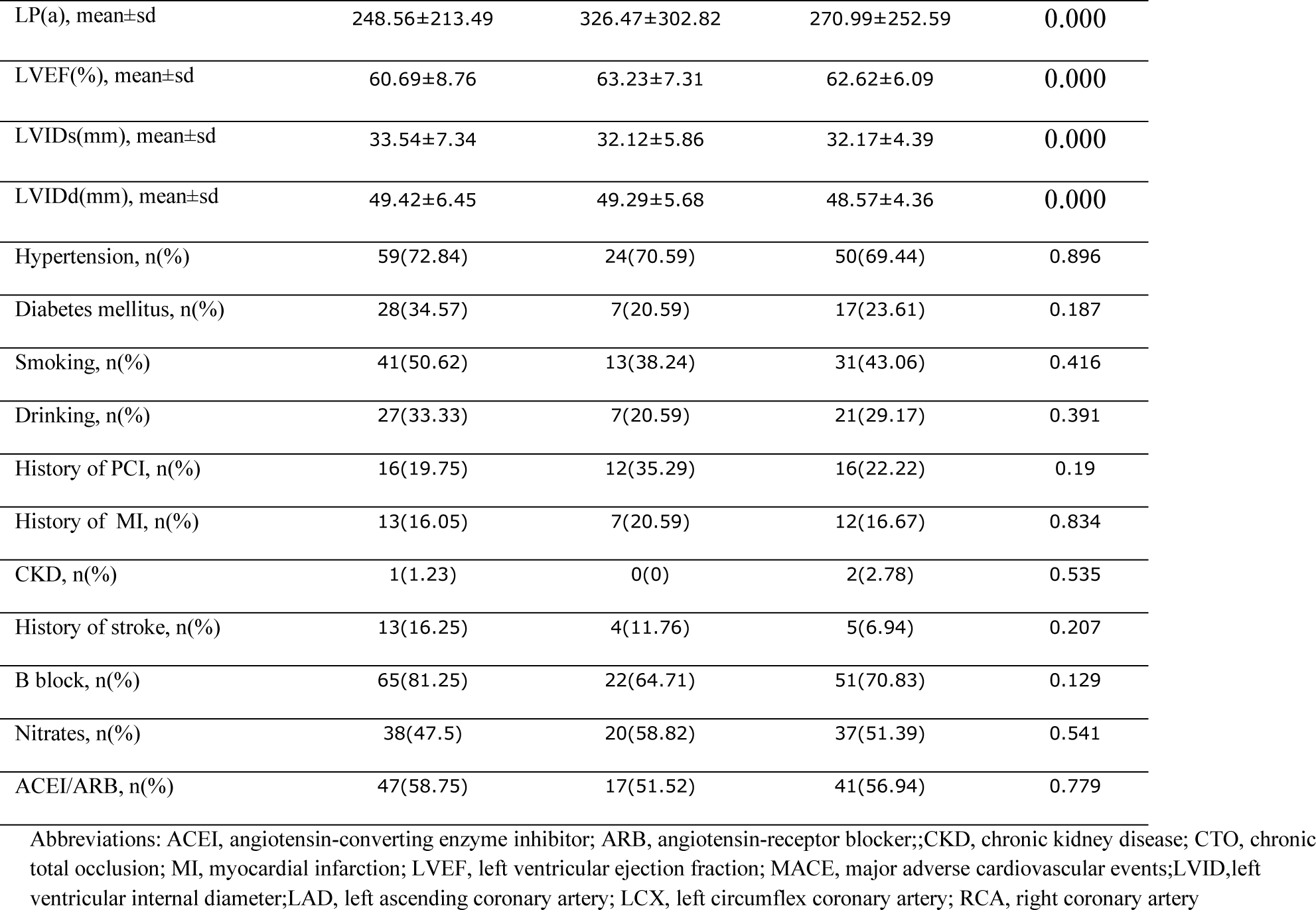
baseline clinical and in-hospital outcome of all patients in the different vessels (LAD, LCX and RCA)

**Table 4.**
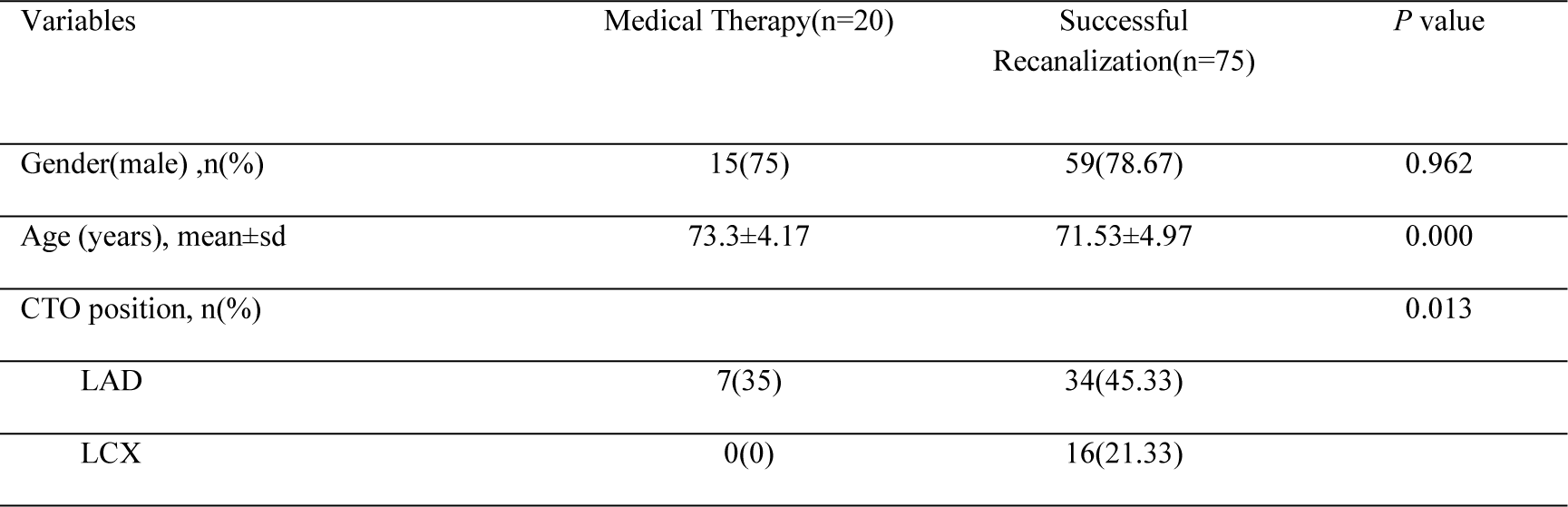
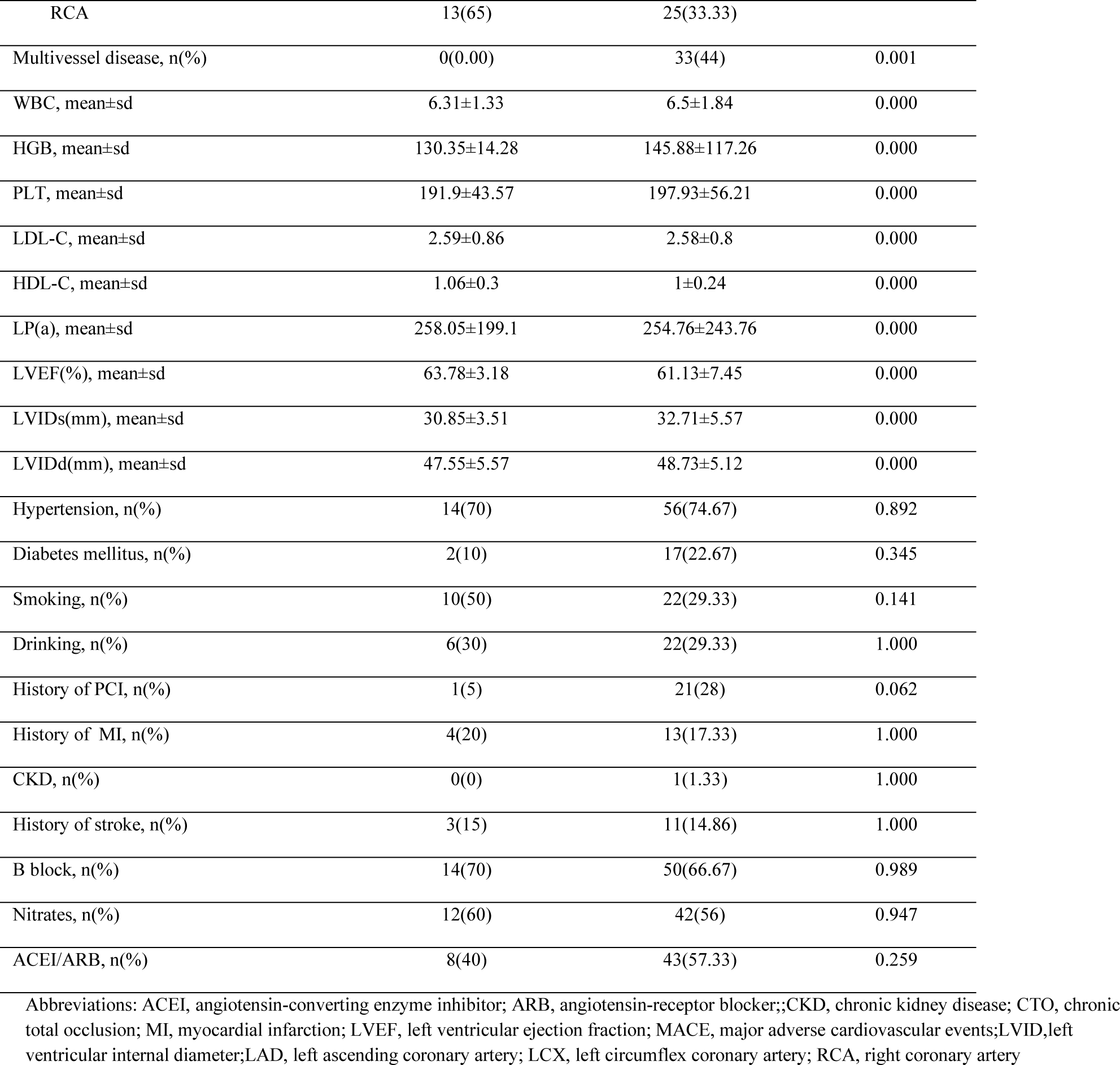
baseline clinical and in-hospital outcome of all patients in the Successful Recanalization and Medical Therapy Groups ≥65 years.

### 3.2 Clinical Outcomes and Kaplan-Meier survival curves

The median duration of follow-up was 78 months. Comparing clinical events between CTO-PCI group and CTO-OMT group, CTO-PCI treatment effect appears to show significant reduction in MACCE(29.55% vs 21.95% p=0.028), mainly contribute to cardiac death (p=0.045) (table 5); Figure 1(a) is the KM curve which demonstrated that it was evident in crossing over at around 125 months, indicating that the data did not meet the proportional hazards assumption. In cases of crossed survival curves, the results of the Log-rank test may be less reliable. Using landmark method, 124 months was taken as the time node. Figure 1(b) demonstrated that CTO-PCI significantly increased the no-event survival rate than the CTO-OMT group in less than 124 months (log-rank test p=0.024). There was no difference in all-cause mortality between the 2 groups (Figure 1c) (log-rank test, p = 0.93). Figure 2 showed that MACCE incidence between single vessel CTO lesions and single vessel CTO combined with multiple vessel lesions were parallel (log-rank test p=0.54). Moreover, Figure 3 showed that the cumulative survival of LAD is the highest among different branch lesion groups (log-rank test p=0.044). Furthermore, CTO-PCI treatment effect in elderly showed significant reduction in MACCE (35.00% vs 21.33% p=0.001), mainly contribute to cardiac death (table 6). The no-event survival probability in elderly increased significantly in PCI treatment group than OMT group (log-rank test p=0.008, Figure 4).

**Figure 1(a).**
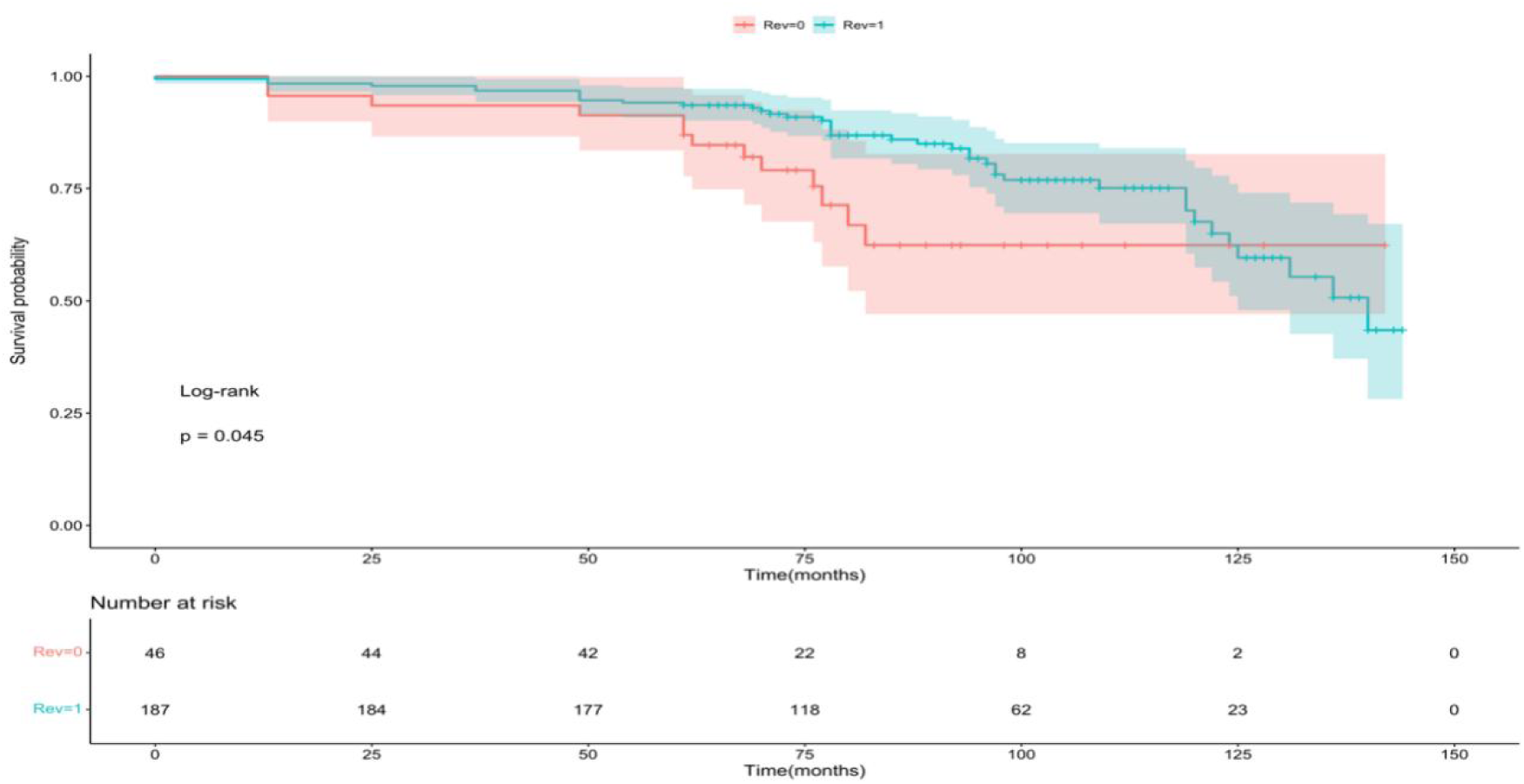
The incidence of major adverse cardiovascular events (including cardiac death, unplanned revascularization, stroke, recurrent myocardial infarction) was compared in the Successful Recanalization and Medical Therapy Groups (Rev0= Medical Therapy Group, Rev1=Successful Recanalization group)

**Figure 1(b).**
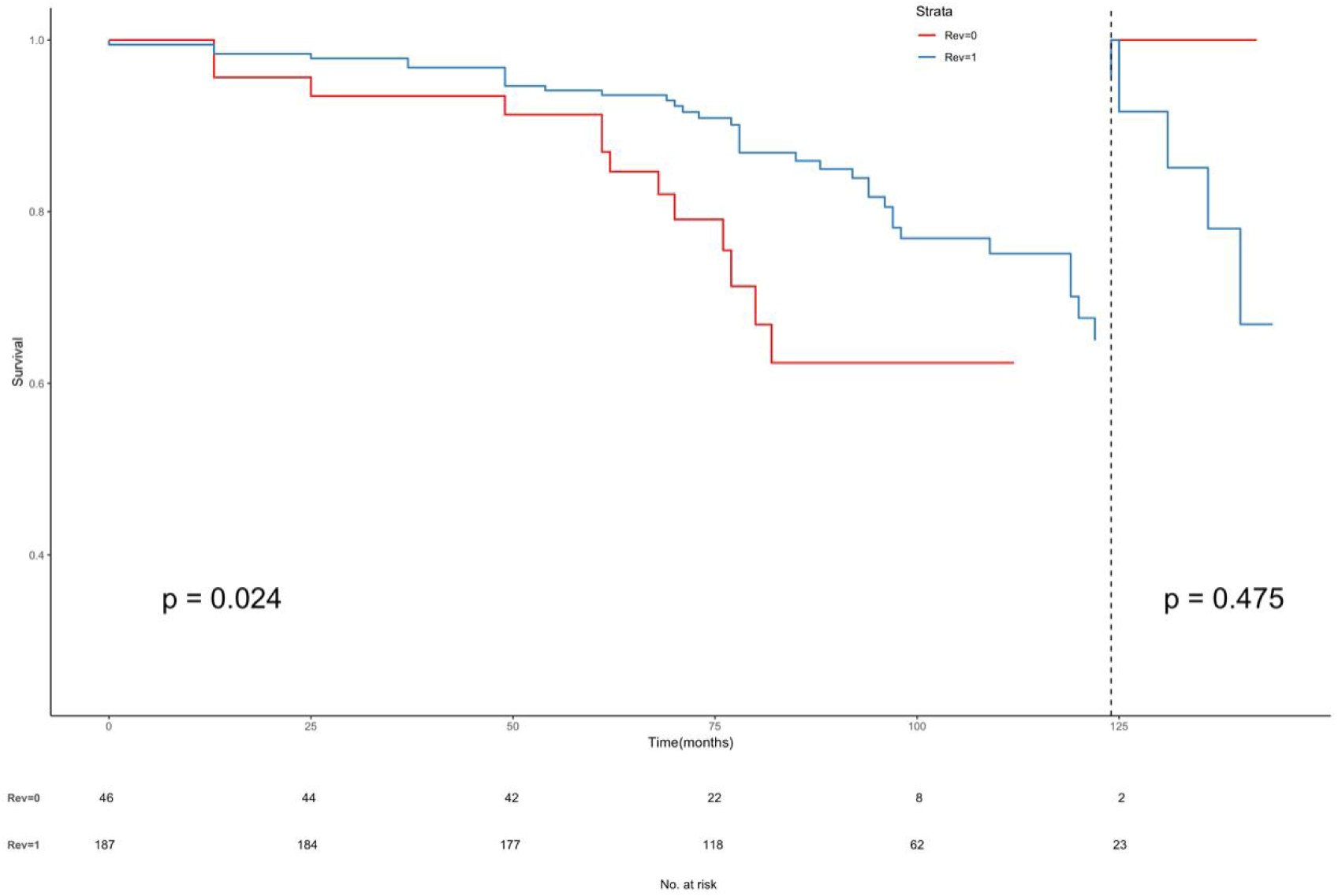
Using landmark method, 124 months was used as the time node (Rev0= Medical Therapy Group, Rev1=Successful Recanalization group)

**Figure 1(c).**
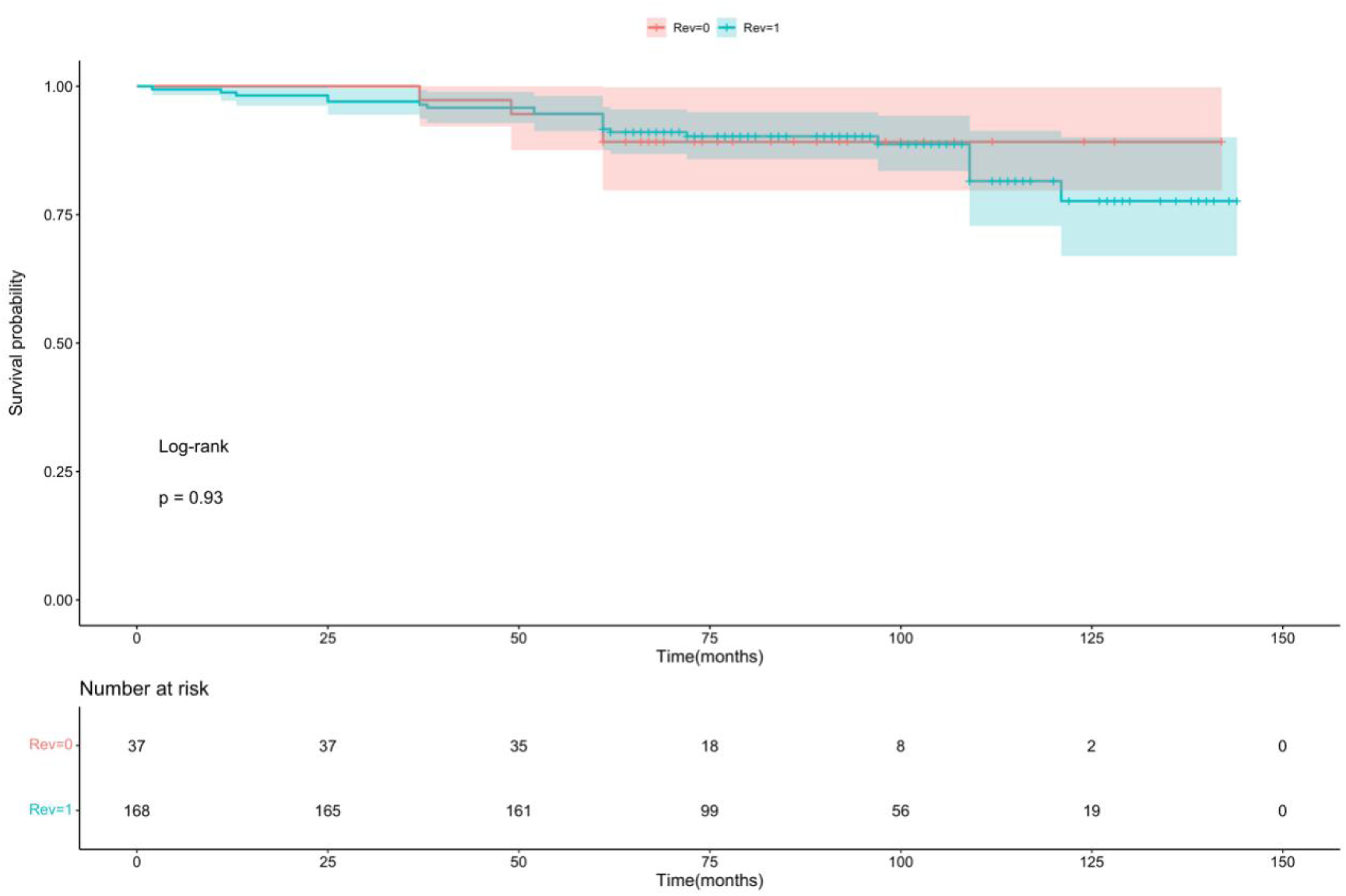
The secondary endpoint - all-cause death (Rev0= Medical Therapy Group, Rev1=Successful Recanalization group)

**Figure 2.**
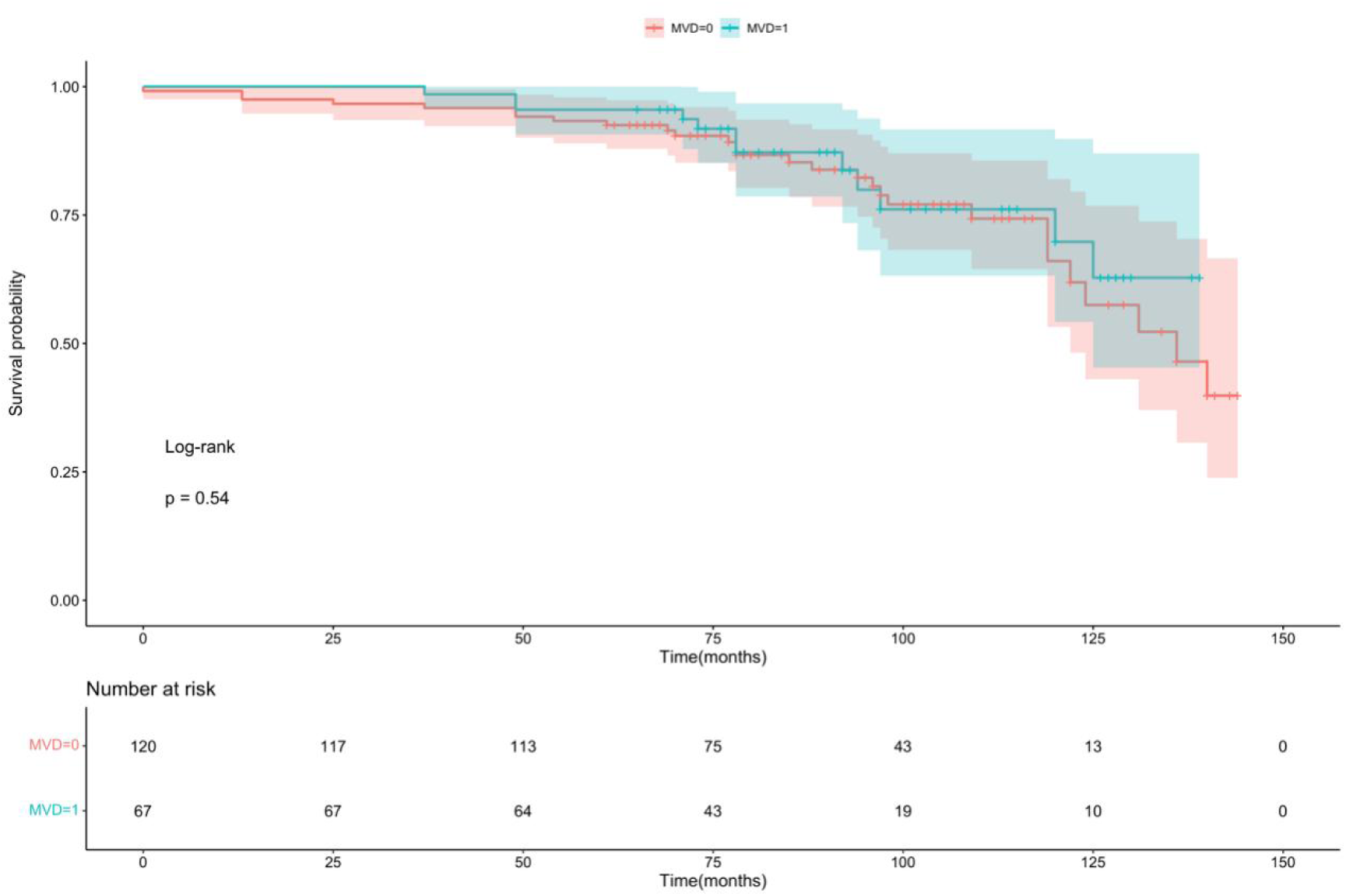
Comparison of the events of CTO combined with multi-vessels and CTO alone groups (MVD0=single-vessel CTO, MVD1= single-vessel CTO combined with multi-vessel lesions)

**Figure 3.**
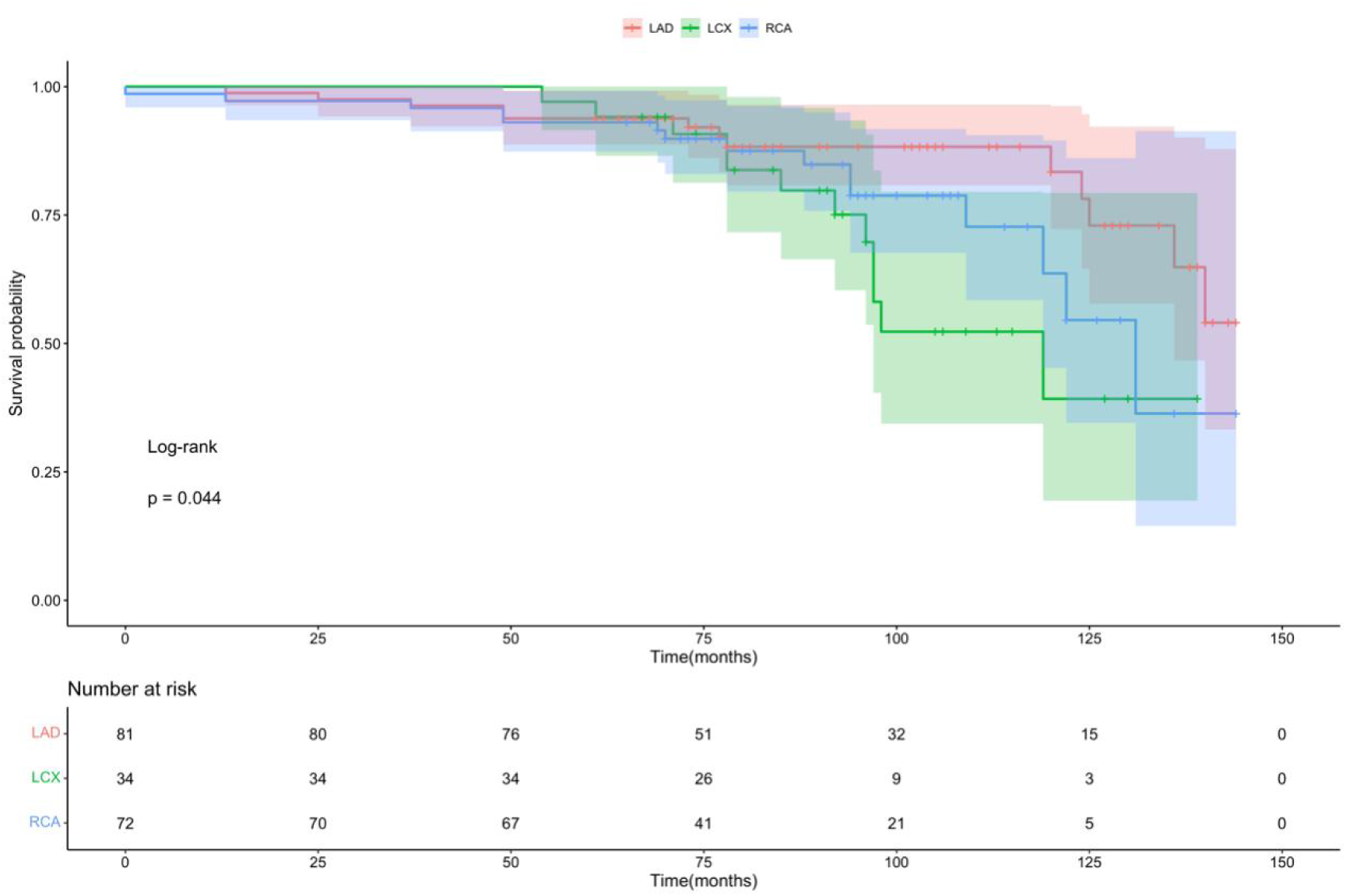
Comparison of the events of CTO in different vessels (LAD, LCX and RCA)

**Figure 4.**
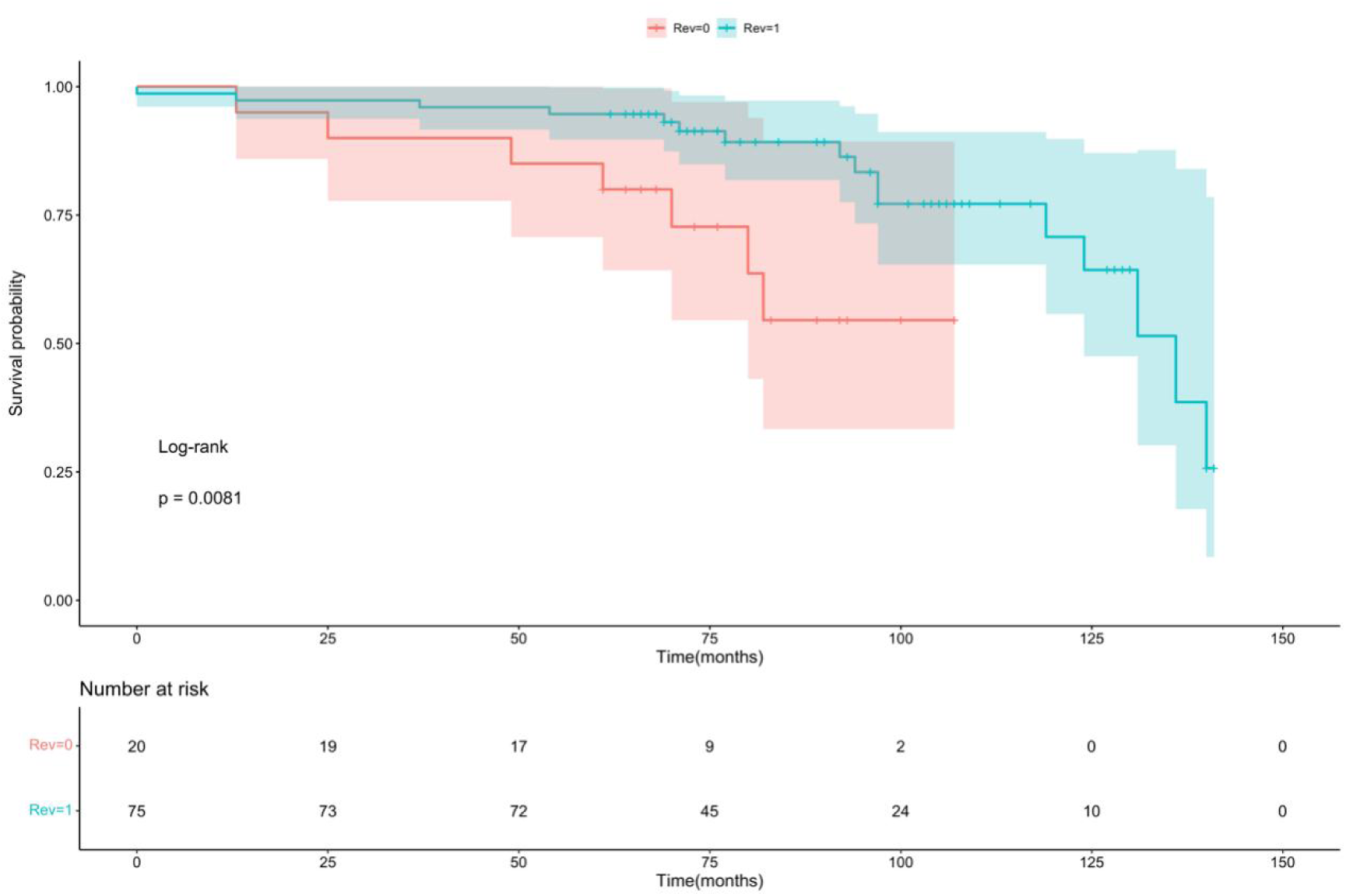
Successful Recanalization and Medical Therapy Groups ≥65 years(Rev0= Medical Therapy Group, Rev1=Successful Recanalization group)

**Table 5.** Clinical Outcomes of All Patients with CTOs in the Successful Recanalization and Medical Therapy Groups.

| Events | Medical Therapy,n(%) | Successful Recanalization,n(%) | Univariable Analysis |  |  | Multivariable-Adjusted |  |  |
| --- | --- | --- | --- | --- | --- | --- | --- | --- |
|  |  |  | HR | 95%CI | p | HR | 95%CI | p |
| MACCE | 13(29.55) | 36(21.95) | 0.6 | (0.32-1.1) | 0.121 | 0.47 | (0.24-0.92) | 0.028 |
| Cardiac death | 7(15.91) | 13(7.93) | 0.6 | (0.3-1.2) | 0.133 | 0.36 | (0.13-0.98) | 0.045 |
| Unplanned revascularizatio | 6(13.64) | 20(12.20) | 0.58 | (0.3-1.1) | 0.113 | 0.53 | (0.21-1.37) | 0.189 |
| Stroke | 0(0.00) | 2(1.22) | - | - | - | - | - | - |
| myocardial infarction | 0(0.00) | 1(0.61) | - | - | - | - | - | - |

**Table 6.** Clinical Outcomes of ≥65 years Patients with CTOs in the Successful Recanalization and Medical Therapy Groups.

| Events | Medical Therapy,n(%) | Successful Recanalization,n(%) | Univariable Analysis |  |  | Multivariable-Adjusted |  |  |
| --- | --- | --- | --- | --- | --- | --- | --- | --- |
|  |  |  | HR | 95%CI | p | HR | 95%CI | p |
| MACCE | 7(35.00) | 16(21.33) | 0.29 | (0.11-0.77) | 0.013 | 0.13 | (0.04-0.44) | 0.001 |
| Cardiac death | 4(20.00) | 6(8.00) | 0.28 | (0.074-1.1) | 0.060 | 0.15 | (0.02-0.91) | 0.040 |
| Unplanned revascularization | 3(15.00) | 9(12.00) | 0.33 | (0.081-1.3) | 0.122 | 0.20 | (0.04-1.14) | 0.071 |
| Stroke | 0(0.00) | 1(1.33) | - | - | - | - | - | - |
| myocardial infarction | 0(0.00) | 0(0.00) | - | - | - | - | - | - |

### 3.3 Multivariate Cox analysis

Further multivariate analysis between two therapy groups, on different revascularization vessels, as well as in elderly patients identified all factors which were associated with MACE occurrence. Table 7 showed that revascularization with ACEI / ARB was beneficial to reduce the incidence of MACE events significantly (Z = -2.201, P = 0.028). Similarly, higher HGB and higher LVEF were associated with statistically significant lower incidence of MACE (P=0.005 and 0.007 respectively).

**Table 7.** Results of the multivariate Cox analysis of MACE events between Successful Recanalization and Medical Therapy Groups.

| Variable | Ratio | HR | 95%CI | Z statistic | P value |
| --- | --- | --- | --- | --- | --- |
| Completion of revascularization | -0.74854 | 0.47 | (0.24-0.92) | -2.201 | 0.028 |
| ACEI/ARB | -0.74826 | 0.47 | (0.26-0.87) | -2.425 | 0.015 |
| HGB | -0.02098 | 0.98 | (0.96-0.99) | -2.801 | 0.005 |
| LVEF | -0.04444 | 0.96 | (0.93-0.99) | -2.720 | 0.007 |

Abbreviations: ACEI, angiotensin-converting enzyme inhibitor; ARB, angiotensin-receptor blocker;;CKD, chronic kidney disease; CTO, chronic total occlusion; MI, myocardial infarction; LVEF, left ventricular ejection fraction; MACE, major adverse cardiovascular events;LVID,left ventricular internal diameter;LAD, left ascending coronary artery; LCX, left circumflex coronary artery; RCA, right coronary artery

Table 8 demonstrated multivariate analysis on three major vessels, wherein males, patients with hypertension, ACEI/ARB or higher LVEF were associated with statistically significant lower incidence of MACE. Higher lipoprotein showed a decreased tendency of MACE with no statistical difference.

**Table 8.** Results of the multivariate Cox analysis of MACE events in the different vessels (LAD, LCX and RCA)

| Variable | Ratio | HR | 95%CI | Z statistic | P value |
| --- | --- | --- | --- | --- | --- |
| Gender | 2.0157498 | 7.51 | (2.54-22.19) | 3.645 | 0.000 |
| CTO | 1.0673795 | 2.91 | (1.11-7.61) | 2.175 | 0.030 |
| hypertension | -1.3717306 | 0.25 | (0.1-0.63) | -2.937 | 0.003 |
| ACEI/ARB | -1.3534238 | 0.26 | (0.09-0.74) | -2.517 | 0.012 |
| LVEF | -0.1151982 | 0.89 | (0.85-0.93) | -4.726 | 2.28e-06 |
| Lipoprotein(a) | -0.0012234 | 1.00 | (1-1) | -1.374 | 0.170 |

Abbreviations: ACEI, angiotensin-converting enzyme inhibitor; ARB, angiotensin-receptor blocker;;CKD, chronic kidney disease; CTO, chronic total occlusion; MI, myocardial infarction; LVEF, left ventricular ejection fraction; MACE, major adverse cardiovascular events;LVID,left ventricular internal diameter;LAD, left ascending coronary artery; LCX, left circumflex coronary artery; RCA, right coronary artery

Furthermore, analysis in elderly showed that complete revascularization reduced the incidence of MACE events in patients and were statistically significant (Z = -3.342, P = 0.001). In addition, LAD opening, β blocker, higher HGB and higher LVEF were associated with significant lower incidence of MACE events (Table 9).

**Table 9.** Results of the multivariate Cox analysis of MACE events in Successful Recanalization and Medical Therapy Groups ≥65 years.

| Variable | Ratio | HR | 95%CI | Z statistic | P value |
| --- | --- | --- | --- | --- | --- |
| Completion of revascularization | -2.01191 | 0.13 | (0.04-0.44) | -3.342 | 0.001 |
| CTO(LAD) | -1.07851 | 0.34 | (0.12-0.94) | -2.077 | 0.038 |
| B-block | -1.24943 | 0.29 | (0.12-0.71) | -2.688 | 0.007 |
| HGB | -0.03521 | 0.97 | (0.94-0.99) | -3.183 | 0.001 |
| LVEF | -0.07873 | 0.92 | (0.88-0.97) | -3.108 | 0.002 |
| HDL-C | 2.17829 | 8.83 | (2.06-37.91) | 2.930 | 0.003 |

Abbreviations: ACEI, angiotensin-converting enzyme inhibitor; ARB, angiotensin-receptor blocker;;CKD, chronic kidney disease; CTO, chronic total occlusion; MI, myocardial infarction; LVEF, left ventricular ejection fraction; MACE, major adverse cardiovascular events;LVID,left ventricular internal diameter;LAD, left ascending coronary artery; LCX, left circumflex coronary artery; RCA, right coronary artery

## 4. Discussion

To date, the clinical efficacy of CTO-PCI revascularization is still controversial. A previous multi-center study [18] showed that successful of CTO-PCI reduced the risk of long-term death compared with failed procedures (8.86 vs 16.8%, P<0.0001). Another study reported that, after a median follow-up time of 66 months, failure of CTO-PCI could significantly increase the incidence of MACE (major adverse cardiovascular events) by about half compared with successful revascularization (P = 0.002) [20]. A meta-analysis of 25 previous studies [21] also revealed that CTO-PCI significantly reduced mortality (OR = 0.52), angina symptoms (OR = 0.38), stroke risk (OR = 0.72), and consequent CABG procedures (OR = 0.18) compared with CTO-OMT. Moreover, several observational studies and meta-analysis have revealed favorable effects on CTO-PCI treatment, including relieved angina and improved quality of life [22,23], potentially enhances myocardial electrical stability, and improved LV systolic function [24]. In contrast, Some studies reported that successful CTO-PCI did not improve the long-term survival [13,14].

Two well-known randomized controlled trials systematically compared CTO-PCI with no CTO-PCI. In DECISION-CTO trial, 834 patients were randomly assigned to the CTO-PCI (n=417) or no CTO-PCI (n=398) group. After a median follow-up of 4 years, there was no significant difference in the incidence of cardiac death, myocardial infarction, stroke, and target vessel revascularization between the CTO-PCI and no CTO-PCI group [25]. However, the power of this trial was limited by the large number of protocol violations, higher crossover rate along with slow enrollment, which may lead to an underestimate of CTO-PCI strategy. EUROCTO trial enrolled 396 patients comparing PCI and OMT in CTO patients. The major adverse cardiovascular event rate was parallel in two groups at 12 months. The incidence of cardiovascular death, or nonfatal myocardial infarction between PCI and OMT was not significantly different at 3 years [26]. Overall, both DECISION-CTO and EUROCTO were not able to conclude CTO therapy strategies due to enrollment size, follow-up duration as well as inconsistent enrollment criteria. Therefore, the efficacy and safety of CTO-PCI deserve further investigation.

Our study demonstrated that successful CTO-PCI significantly decreased long-term MACCE and cardiac death compared to CTO-OMT. Therefore, based on our study, CTO-PCI is the effective strategy for reduction of MACCE and cardiac death. In addition, MACCE and cardiac death in elderly group consistently showed the beneficial effects of PCI. MACCE in CTO vessel with or without multi-vessel occlusions were parallel, partially because our enrollment size was not large enough to statistically analyze. Moreover, revascularization of LAD had higher no-event survival probability. In summary, our study is characteristic of: (1) Enrollment included both patients with single-vessel CTO lesions and single-vessel CTO combined with multi-vessel lesions; (2) We studied CTO therapy in patients with single-vessel CTO lesions and patients with multi-vessel lesions; (3)CTO lesions in LAD, LCX, or RCA and therapies effects were separately investigated; (4) CTO therapy in elderly was further investigated. The rationale of the study design was based on: (1) the occluded vessel has been opened after coronary interventional therapy, the ischemic myocardium in the blood supply area is reperfused and revived, thereafter, the symptoms, such as angina pectoris, are reduced; (2)Additional medications could slow down or even reverses part of the ventricular remodeling, so as to further improve the cardiac function and prognosis. Above all, not all CTO lesions need to be opened; (3) Elderly patients have lower physical activities and better tolerance to ischemia, which could offset the benefit of revascularization. For some special populations, such as elderly patients, patients with cancer, or coagulation dysfunction, medications are also necessary alternatives.

## 5. Limitations

This study has some limitations. First of all, it is a single-center retrospective observational study, and there is inevitable selectivity bias, it still needs to be further investigated by large-scale, multi-center prospective studies. Second, the relatively small enrollment in the OMT group and the lower incidence of death and myocardial infarction require a larger enrollment for analysis. Third, due to the rapid development of CTO-PCI technology, along with the experience accumulation of the operators, the success rate of PCI procedures keeps increasing, and operators’ skill at that time may not be equivalent to the current. Finally, patients in this study were not examined by myocardial radionuclide imaging or cardiac magnetic resonance imaging before CTO intervention, and in-depth assessment of ischemia and myocardial viability in the occluded vascular blood supply area is helpful to select patients who can benefit more from CTO-PCI therapy.

## 6. Conclusion

Successful PCI in CTO patients is related to a significant decrease of MACCE compared with OMT. The higher MACCE incidence in the CTO-OMT patients mainly attributed to cardiac death.

## Data Availability

It is availability of all data referred to in the manuscript

## 7. CONFLICT OF INTEREST

The authors declare that they have no competing interests.

## Notes

**FUNDING** **This work was supported by the Foundation of Clinical Medical Research Center of Yili Autonomous Prefecture (yl2023ms01)**

### Competing Interest Statement

The authors have declared no competing interest.

### Clinical Trial

2023-SR-893

### Funding Statement

This work was supported by the Foundation of Clinical Medical Research Center of Yili Autonomous Prefecture (yl2023ms01)

